# Transferable cancer detection from cell-free DNA fragment lengths through extensions of non-negative matrix factorization

**DOI:** 10.1101/2025.09.25.25336628

**Authors:** Ludvig Renbo Olsen, Jakob Qvortrup Holsting, Nicolai Juul Birkbak, Lars Dyrskjøt, Jakob Skou Pedersen, Claus Lindbjerg Andersen, Søren Besenbacher

## Abstract

**Motivation:** The fragment length distribution of cell-free DNA (cfDNA) can reveal the presence of circulating tumor DNA (ctDNA) in a plasma sample. A previous study documented that non-negative matrix factorization (NMF) can extract relevant features from the fragment length distributions. These distributions, however, are affected by technical biases. When NMF is performed on samples from multiple datasets, some of the extracted signatures often capture these technical biases rather than the actual biological differences.

**Results:** We present two methods for extracting biologically meaningful NMF signatures across heterogeneous datasets with varying technical biases. Using simulated data, we first demonstrate that these new methods are more effective at estimating the true proportions of the underlying processes. We then show that the methods increase the transferability of fragment length signatures to cfDNA datasets from external labs. Classification models using the two proposed NMF extensions achieve an average cross-dataset AUC of 0.842 and 0.814, compared to 0.776 for standard NMF and 0.688 for a set of previously reported manually selected fragment length features. We further show that one of the proposed methods requires only 1-5 samples to estimate the batch effect during inference.

**Availability and Implementation:** The source code and instructions on how to run it are available at: https://github.com/BesenbacherLab/batch-NMF

## 1 Introduction

Analyzing circulating cell-free DNA (cfDNA) from the blood offers a minimally invasive strategy for cancer diagnosis. Distinguishing the circulating tumor DNA (ctDNA) fragments from the much larger pool of healthy cfDNA fragments is, however, challenging, as only a small subset of ctDNA fragments carry mutations that clearly mark their origin. Analyzing cfDNA fragment lengths is a promising mutation-independent strategy to detect the presence of cancer from the distinctive length distribution of ctDNA.

Early studies used gel electrophoresis (Jahr et al. 2001; Giacona et al. 1998) or qPCR (B. G. Wang et al. 2003) to compare the length distribution of cfDNA in cancer patients with that of healthy individuals. Later studies have used whole-genome sequencing (WGS) to obtain a more detailed measurement of the cfDNA length distribution (Shi et al. 2020). Jiang et al. (Jiang et al. 2015) used WGS to compare the cfDNA lengths in chromosome arms with tumor amplifications to the length distribution in chromosome arms with tumor deletions and concluded that ctDNA had a higher proportion of short (<150bp) fragments. Besides this general shortening, Mouliere et al. (Mouliere et al. 2018), 2018) also reported that cancer patients exhibited more pronounced oscillations with a 10 bp periodicity in short fragments.

The size distribution of a cfDNA sample is not solely determined by whether it comes from a cancer patient. Recent studies have found that multiple factors can affect the fragment length distribution, including DNA methylation level (Zhou et al. 2022), the presence of histone modifications (Bai et al. 2024), and the abundance of different DNA-cleaving enzymes (Serpas et al. 2019; Han et al. 2020). It is thus likely that there are cancer type-specific and individual differences in the fragment length distribution, even among cancer patients that have the same tumor fraction (Thierry 2023).

If multiple fragmentation processes are involved in forming the cfDNA fragments in a sample, it would be useful to simultaneously learn the characteristics of these processes and their importance in different samples. Renaud et al. suggested using non-negative matrix factorization (NMF) to achieve this (Renaud et al. 2022).

A general problem for analyses of cfDNA fragment length is that the length distributions are affected by both pre-analytical (choice of kit for extraction, library preparation, purification, and amplification) (Markus et al. 2018; Chang et al. 2021; Dabney and Meyer 2012) and analytical (type of sequencing machine and bioinformatic handling) (Gohl et al. 2019; H. Wang et al. 2025) factors. Since these confounding factors are usually constant within a dataset, they will not significantly affect the ability to distinguish cases from controls within that dataset. Such differences in sample processing or sequencing technology between datasets will, however, mean that a method trained on one dataset cannot be expected to generalize to another dataset. Technical differences between datasets also complicate training classification methods on samples from multiple datasets, as confounding factors may obscure the true case-control differences, increasing the risk of mistaking technical variation for true biological signals in unbalanced data.

In this article, we propose two extensions of NMF that incorporate possible confounding factors due to technical differences between datasets during the signature discovery. *Batch NMF* (bNMF) is a general method for finding NMF signatures in a dataset consisting of multiple batches where the global distribution of features is shifted between batches. *Sigmoid Filter NMF* (sfNMF) is particularly suited to analyze fragment length data and assumes that the differences between batches can be described as a combination of monotonic increasing filters and monotonic decreasing filters.

## 2 Methods

### 2.1 NMF

Let *X*_*N* × *M*_ be a matrix where the *N* rows are the sum-to-one normalized fragment length histograms of cfDNA samples with *M* different fragment lengths. NMF can then be used to factorize the non-negative matrix, *X*_*N* × *M*_, into two smaller non-negative matrices *W*_*N* × *C*_ and *H*_*C* ×*M*_ (Figure 1b). Where the number of signatures, *C*, is much smaller than *M*, leading to a substantial reduction in the dimensionality of the data. In many situations, non-negativity makes matrix factorizations more interpretable than unconstrained factorization methods, such as principal component analysis (PCA). This is, for instance, the case when analyzing mutational processes where we know that processes can only add mutations, not remove them. Consequently, NMF fits the generating mutational process better than PCA, resulting in signatures that more accurately reflect the true generating processes (Alexandrov et al. 2013).

**Figure 1.**
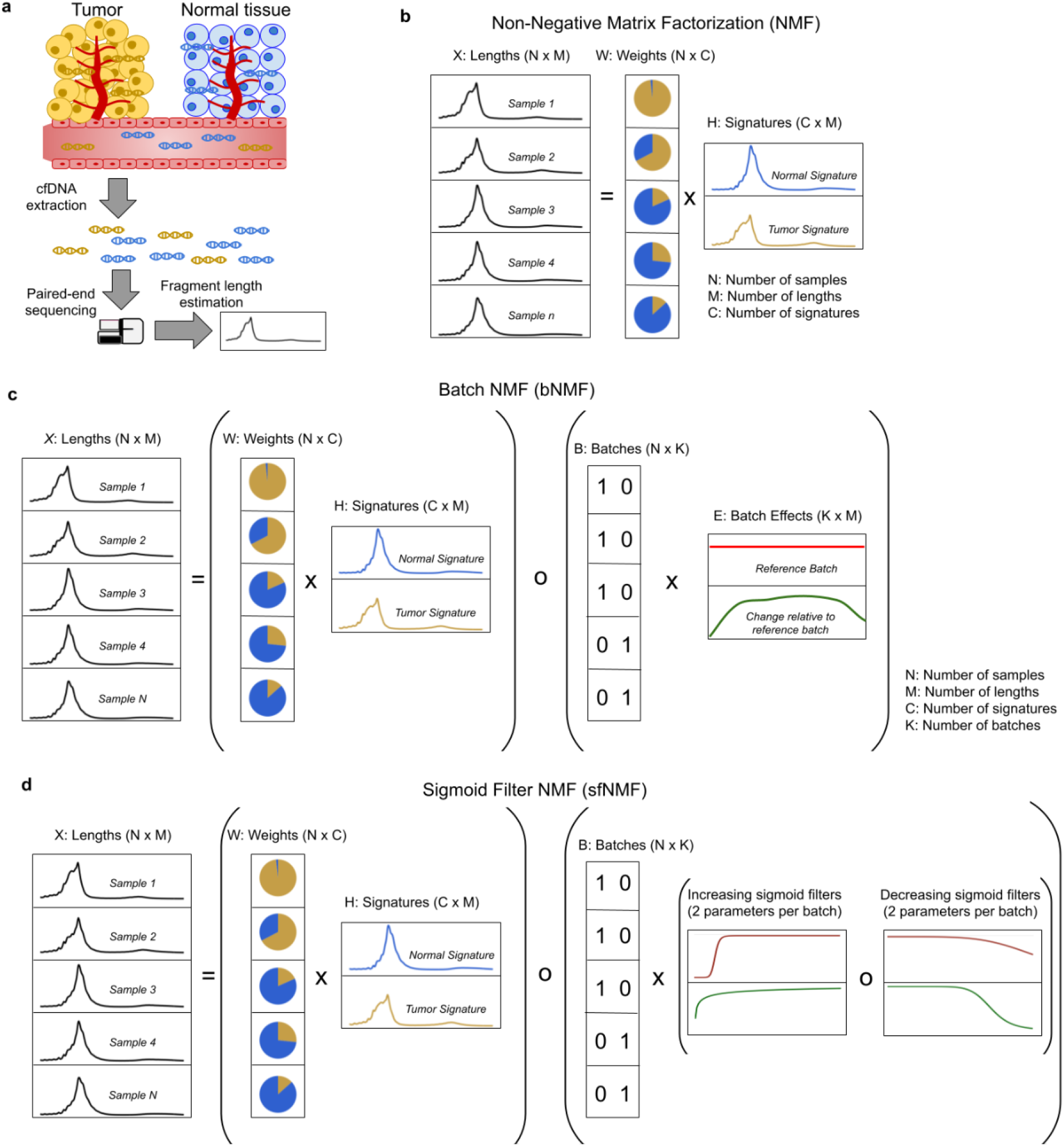
Method overview. **a**. The length of cfDNA fragments can be estimated using paired-end sequencing. **b**. Standard NMF. **c**. The proposed Batch NMF method. “o” denotes the Hadamard (element-wise) matrix product. **d**. The proposed Sigmoid Filter NMF method. Adapted from figures in (Renaud et al. 2022).

For a given *C*, NMF will find the optimal matrices, *W*_*N* × *C*_ and *H*_*C* × *M*_, so that *W* × *H* approximates *X*_*N* × *M*_ in terms of some distance measure *D*:

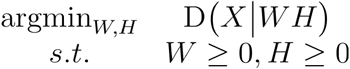

The distance function *D* (*X* │ *W H*) is typically the Frobenius norm or the Kullback-Leibler (KL) divergence, and it is usually minimized using the very efficient multiplicative update algorithm (Lee and Seung 2000).

### 2.2 Batch NMF (bNMF)

When we have samples from *K* different datasets (or batches) that have been processed differently, the NMF signatures will likely capture these technical differences rather than the true biological signals. To avoid this, we propose an extension to NMF called bNMF, where we select one dataset as a reference dataset and then have a matrix specifying how each of the other datasets diverges from it. Let *B* _*N × K*_ be a matrix that one-hot encodes which dataset a sample comes from. The matrix *E*_*K* × *M*_ then estimates the dataset differences (See Figure 1c). We define the bNMF problem as finding the best *W,H*, and *E* that can recreate the input data:

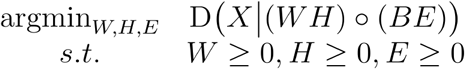

To train this method, we use transformations of unconstrained matrices to turn the non-negative optimization problem into an unconstrained optimization problem:

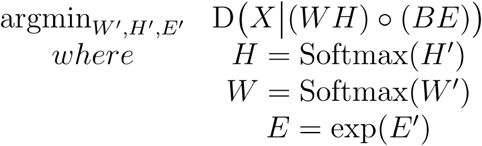

We then fit this unconstrained optimization problem using PyTorch. Using the row-wise softmax function to transform the *H* and *W* matrices ensures that all values in these matrices are positive and that the rows sum to 1. Using the exponential function to transform *E* ensures that all values in this matrix are positive.

### 2.3 Sigmoid Filter NMF (sfNMF)

The bNMF method is a general solution for handling batch effects in non-negative matrix factorization (NMF) models. In our specific application of NMF to cfDNA fragment lengths, we have some knowledge about the processes that cause differences between datasets. Not all the DNA fragments in a given plasma sample will be present in the sequencing data. Some fragments will be lost at different steps, from DNA extraction to library preparation and sequencing. These different steps can be viewed as filters that remove certain fragments that will not be included in the final data. Sometimes, these filters can be biased and preferentially remove either short or long fragments. One source of differences between datasets or batches is the method used to isolate and purify cfDNA (Chang et al. 2021). If magnetic beads are used for one or more steps during library preparation, the number of beads makes a significant difference. A low bead-to-DNA ratio will result in the loss of short fragments from the sample (Rodrigue et al. 2010). The effect of bead purification can thus be described using a monotonically increasing function. Sequencing machines, on the other hand, exhibit varying degrees of bias towards shorter fragments, leading to the loss of long fragments (Gohl et al. 2019). This effect can thus be described using a monotonically decreasing function. To limit the number of parameters and avoid overfitting, we propose an alternative NMF model that fits an increasing sigmoid filter and a decreasing sigmoid filter for each dataset (Figure 1d). As shown in Figure 2, a sigmoid function ranging from 0 to 1 can be described with two parameters,*h* and *n*. Where *h* describes the point where the function takes the value of 0.5, and *n* is a parameter that controls the steepness. If *n* is negative, the function will be an increasing sigmoid (Fig. 2a); otherwise, it will decrease (Fig. 2b).

**Figure 2.**
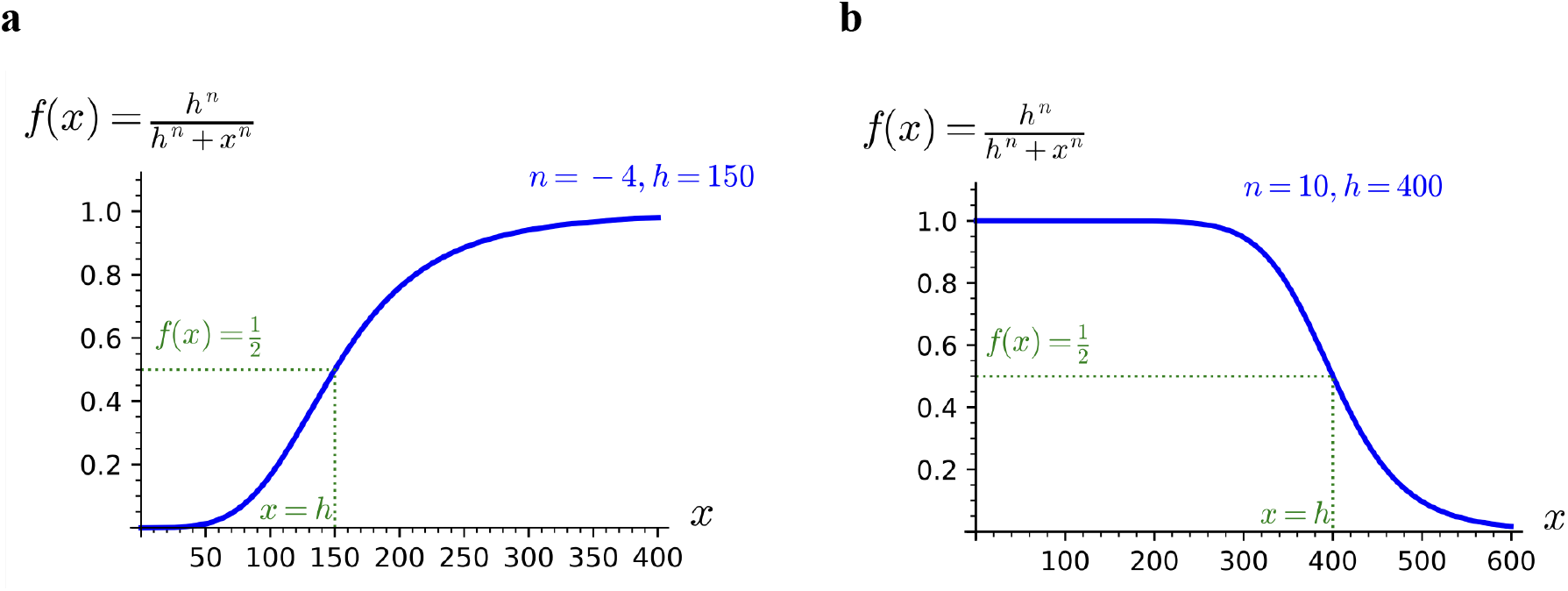
Sigmoid filters. **a**. Example of increasing sigmoid function.*x* is a particular fragment length, and the value of *f* (*x*) describes what fraction of a given length is left after some process has filtered the lengths. **b**. Example of a decreasing sigmoid function. A positive *n* results in a declining sigmoid, while a negative *n* leads to an increasing sigmoid.

Let *L*_*M*_ be a vector of the possible fragment lengths. We then define a sigmoid function that takes two vectors of length *K* and calculates a *M* × *K* matrix, describing the fraction of a given length retained in each batch after the filter has been applied.

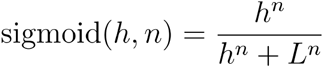

To describe the processes that can remove both short and long fragments, we use four vectors of length *K* called *h*_*i*_,*n*_*i*_, *h*_*d*_, *n*_*d*_ describing the two *increasing* sigmoid and two *decreasing* sigmoid parameters for each dataset. We can then formulate the sfNMF problem as the following optimization task:

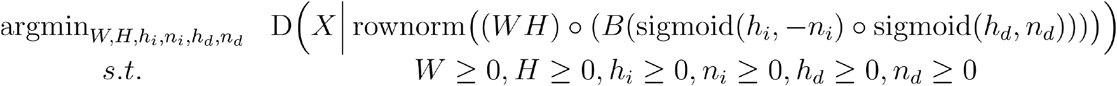

Like bNMF, we also fit this model using PyTorch, applying transformations to ensure the non-negativity constraints are met. We use the softmax transformation to ensure that *W* and *H* are non-negative and their rows sum to one. For *h*_*i*_, *n*_*i*_, *h*_*d*_, and *n*_*d*_, and, we use the softplus transformation to ensure non-negativity.

### 2.4 Test datasets

We included three cfDNA WGS datasets from different labs to test our method in a scenario with high technical variation between datasets (Supp. Table 1). The datasets contained 10 different cancer types, with only colorectal and lung cancer being present in multiple datasets. The sequencing protocols varied between studies, with different library preparation kits and sequencing machines employed. As seen in Figure 3, these differences in sequencing protocols result in differences in the average fragment length distribution in the controls from each dataset.

**Figure 3:**
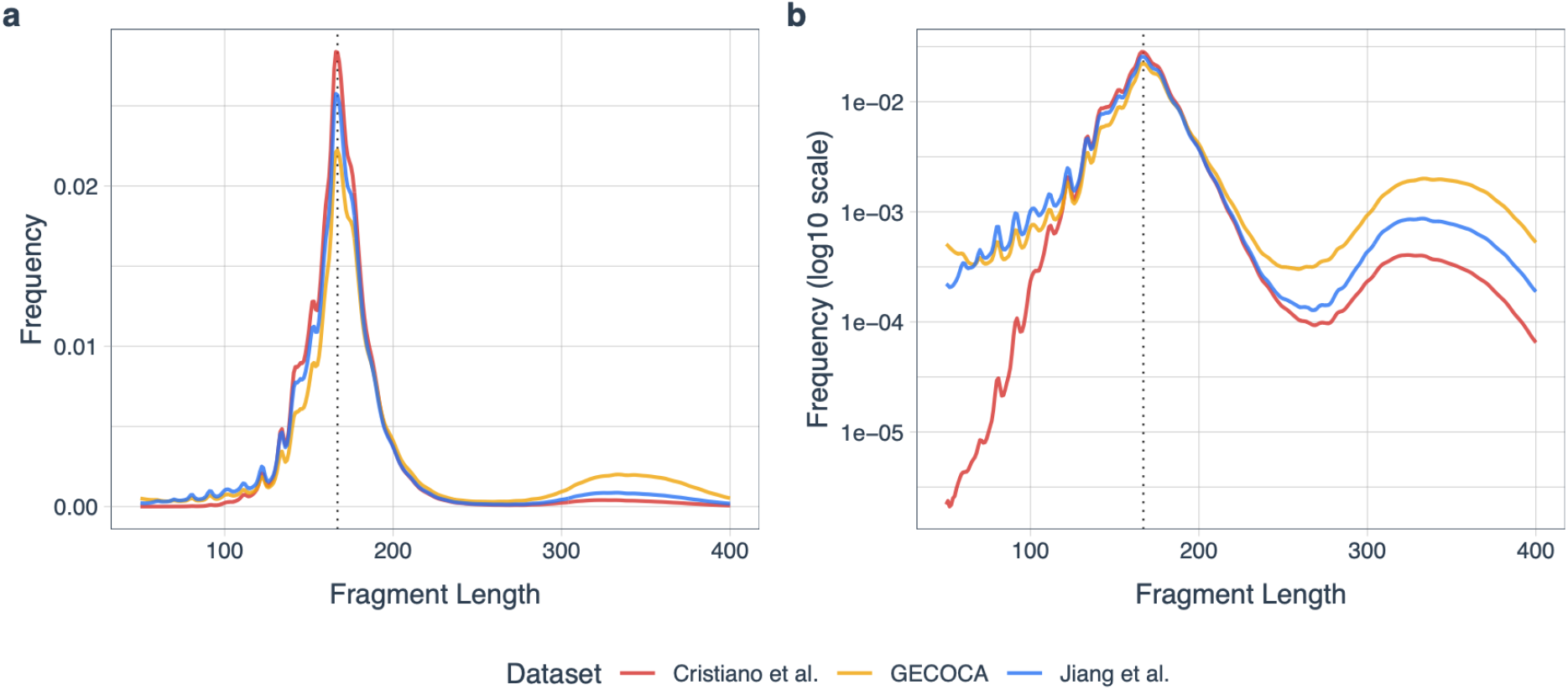
Average fragment lengths for the controls of each dataset. **a**. Average frequency of each fragment length among controls. **b**. Same as **a** but on a log-10 scale.

The GECOCA dataset (Olsen et al. 2024) consisted of 134 patients with colorectal cancer and 34 non-cancer controls. Sequencing libraries were prepared using the KAPA HyperPrep library preparation kit and sequenced on Illumina NovaSeq.

The dataset from Cristiano et al. (Cristiano et al. 2019) contained 230 cancer patients with one of 7 cancer types (54 breast, 35 lung, 34 pancreatic, 28 ovarian, 27 colorectal, 27 gastric, and 25 bile duct). Sequencing libraries were prepared using the NEBNext DNA library preparation kit and sequenced on an Illumina HiSeq 2000 or 2500.

The dataset from Jiang et al. (Jiang et al. 2015) contained 74 cancer patients with one of 5 cancer types (34 hepatocellular carcinoma, 10 colorectal, 10 lung, 10 head and neck squamous cell carcinoma, and 10 nasopharyngeal carcinoma). Libraries were prepared using the KAPA library preparation kit and sequenced on Illumina HiSeq 2000.

### 2.5 Leave-one-dataset-out cross-validation

To test the methods on the three test datasets, we perform leave-one-dataset-out cross-validation. For each of the three datasets, we conduct an experiment where we train both the NMF method and subsequently a LASSO logistic regression model on the other two datasets. We then infer the weight of the signatures found in the training data for each sample in the test data and use the fitted classifier to predict their cancer status. Figure 4 shows an overview of the steps involved in each experiment.

**Figure 4.**
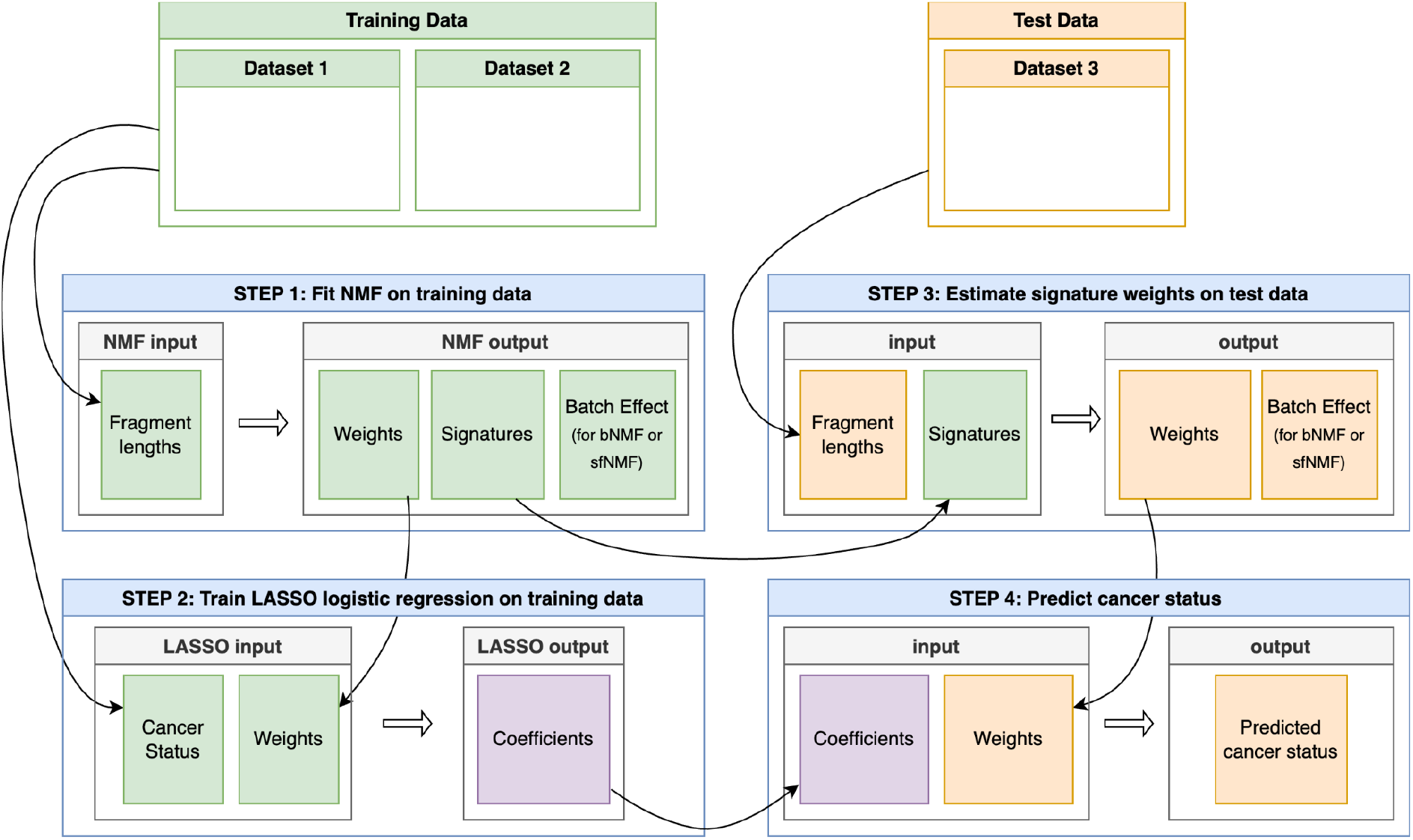
The steps performed for each fold (dataset) in the leave-one-dataset-out cross-validation.

### 2.6 Calculating read length distributions

For each dataset, we first used *cutadapt* to remove any adaptor sequences. The reads were then mapped to hg38 using *bwa-mem*, and duplicates were marked with *picard*. We excluded reads from the genomic regions listed in the ENCODE blacklist v2 (Amemiya, Kundaje, and Boyle 2019) and umap k100 (Karimzadeh et al. 2018) exclusion file. Finally, we counted the fragment lengths of all read pairs that mapped to an autosome with a mapping quality of at least 40 using *ctDNAtool* (https://github.com/BesenbacherLab/ctDNAtool). For the analyses, we only considered fragment lengths between 50 and 400 bp and normalized the counts within each sample so that they summed to 1.

### 2.7 Fitting NMF models

The bNMF and sfNMF models were implemented in PyTorch and fitted using the Adam optimizer. For each run, we tried 10 different random initializations and ran for 40,000 iterations, picking the one with the lowest loss. We continued fitting this model while lowering the learning rate using the *ReduceLROnPlateau* method. If the loss had not improved for 5000 iterations, the learning rate was reduced by 10% until it dropped below 1e-5.

Given a set of signatures found in a training dataset, we can determine the weights of those in a test dataset by fitting a model where the signature matrix, *H*, is fixed and only the remaining parameters (*W* and batch effect parameters) are fitted.

To compare our methods with standard NMF, we also performed decomposition of the normalized input matrix using *scikit-learn* (*sklearn*.*decomposition*.*NMF*) with random initialization, multiplicative updates, and the Kullback-Leibler loss function. To avoid problems with random initialization, the NMF was repeated 10 times with different random initializations, and the fit with the lowest loss was selected. The rows of the estimated weight matrix were subsequently scaled so that they sum to one.

### 2.8 Fitting classification models

Using the weights of the fragment length signatures from each of the three NMF models separately, we used LASSO logistic regression to classify the cancer status of the samples. The classification models were implemented in R using the *tidymodels* and *glmnet* packages. The LASSO regularisation parameter was fitted using 5-fold cross-validation on the training data.

## 3 Results

### 3.1 Results on simulated data

To test the implementations of the two proposed methods, we first ran the methods on a simple simulated dataset. To create this dataset, we mixed two different fragment length signatures with various mixture proportions and applied a filter that removed a fraction of the short and long fragments in half of the samples (Fig. 5a). We then used NMF, bNMF, and sfNMF to estimate the mixture proportions (weights) that had been used to construct the length distributions. Across ten different runs, both bNMF and sfNMF could consistently infer signatures and weights that correlated well with the true values (Fig. 5b). In contrast, standard NMF found weights that had significantly lower correlations with the true weights.

**Figure 5.**
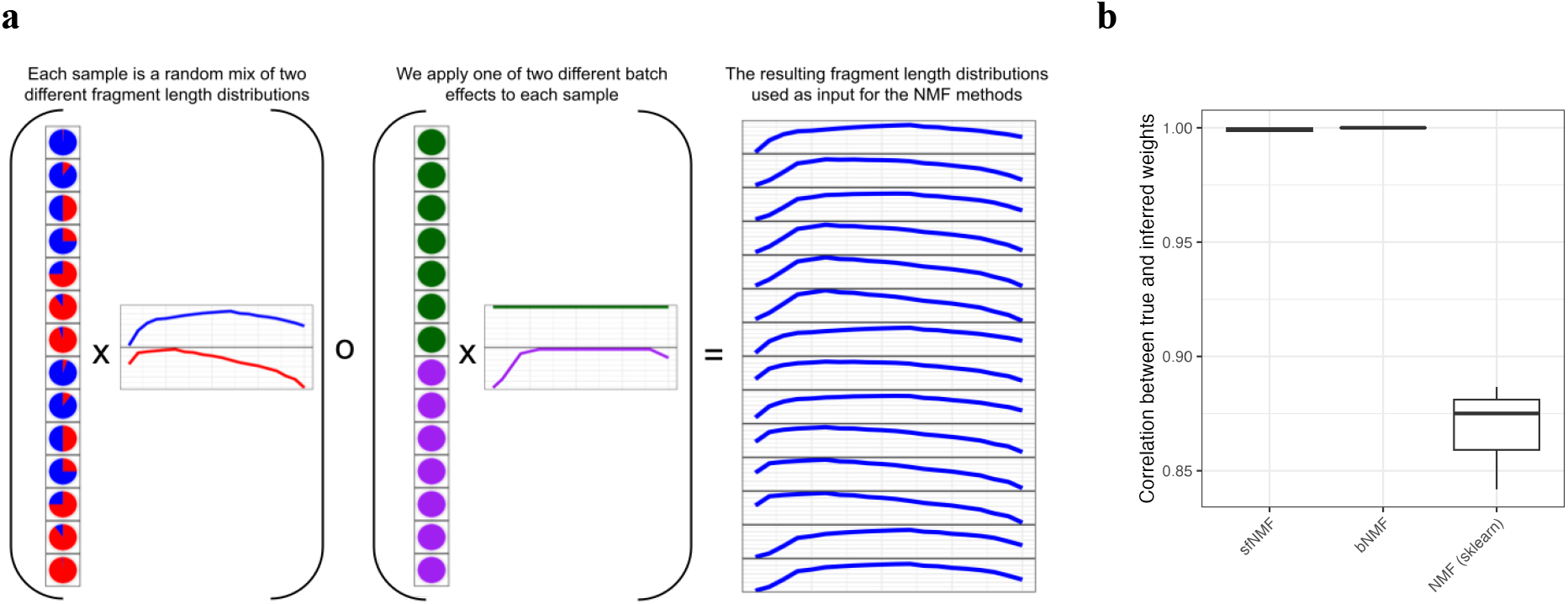
**a**. Simulated data. 14 samples are simulated by mixing two length signatures (shown in red and blue). The first seven are simulated to come from a dataset with no bias in which lengths are observed (green). In the last seven samples, a fraction of short and long fragments are lost (purple). The resulting matrix (blue) is normalized so that each row sums to 1, representing the simulated fragment length distribution for each of the 14 samples. **b** The correlations between the true (simulated) weight matrix and the weight matrices obtained by running NMF, bNMF, and sfNMF on the fragment length distribution matrix.

### 3.2 Predicting cancer status in new datasets

Next, we wanted to test how well a classification method that uses cfDNA fragment length signatures to predict cancer status can generalize to new datasets not encountered during training. To test this, we collected three datasets produced in different labs using different protocols. We then ran leave-one-dataset-out cross-validation experiments, where two of the three datasets were used for training, and the last dataset was used for testing. For each train/test split, we fitted an NMF model on the training data and then trained a LASSO logistic regression classifier to predict cancer status from the weights of the found fragment length signatures. We then estimated the weight of the (fixed) signatures in the test dataset and used the trained classifier to predict cancer status. We use the Area Under the ROC Curve (AUC) to quantify the performance of the models on the test dataset. For two of the datasets, the extended NMF models result in vastly better classification models than standard NMF, while the results are similar for the last dataset (Fig. 6). The performance of the bNMF and sfNMF is, on average, 10% and 9.8% better, respectively, than standard NMF across datasets and different numbers of signatures. For 20 signatures, the average AUC is 0.776 for NMF, 0.842 for bNMF, and 0.814 for sfNMF. For comparison, we trained a logistic regression model on the standardized fragment length proportion features proposed by Mouliere et al. (Mouliere et al. 2018). If P(x) denotes the proportion of fragments in a given length interval x then these features can be described as: P(20 to 150), P(100 to 150), P(160 to 180), P(180 to 220), P(250 to 320), P(20 to 150) / P(160 to 180), P(100 to 150) / P(163 to 169), P(20 to 150) / P(180 to 220), and 10bp amplitudes below 150bp. This achieved an average leave-one-dataset-out cross-validated AUC of 0.688 (GECOCA=0.669, Cristiano et al.=0.731, Jiang et al.=0.663).

**Figure 6.**
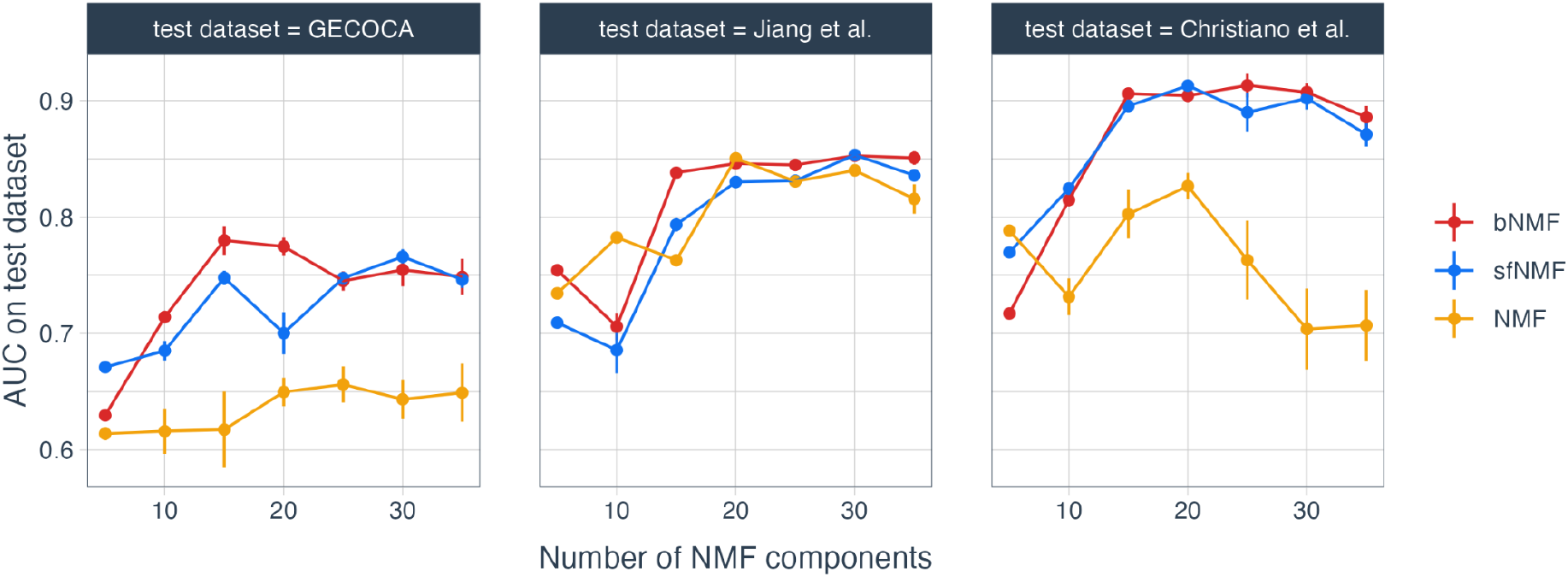
Results from leave-one-dataset-out cross-validation. Each panel shows the area under the ROC curve on a test dataset. The test dataset was neither used during the fitting of the NMF signatures nor the subsequent training of the LASSO logistic regression classifier. Results are shown for different numbers of signatures. The error bars are standard errors of the mean calculated from 10 repeats of the experiment.

A disadvantage of the bNMF model is that it requires estimating a significantly larger number of parameters on the test data than NMF and sfNMF (one per fragment length per batch for bNMF, compared to four per batch for sfNMF). We thus expect it to perform worse when we want to estimate the weight parameters for a small number of test samples. To test this expectation, we divided the test datasets into smaller batches and estimated the weight of 20 signatures found in the training data independently for each of these batches. The results shown in Figure 7 show that bNMF does, in fact, perform worse than sfNMF when we infer the weights independently in smaller batches instead of analysing all test samples jointly.

**Figure 7.**
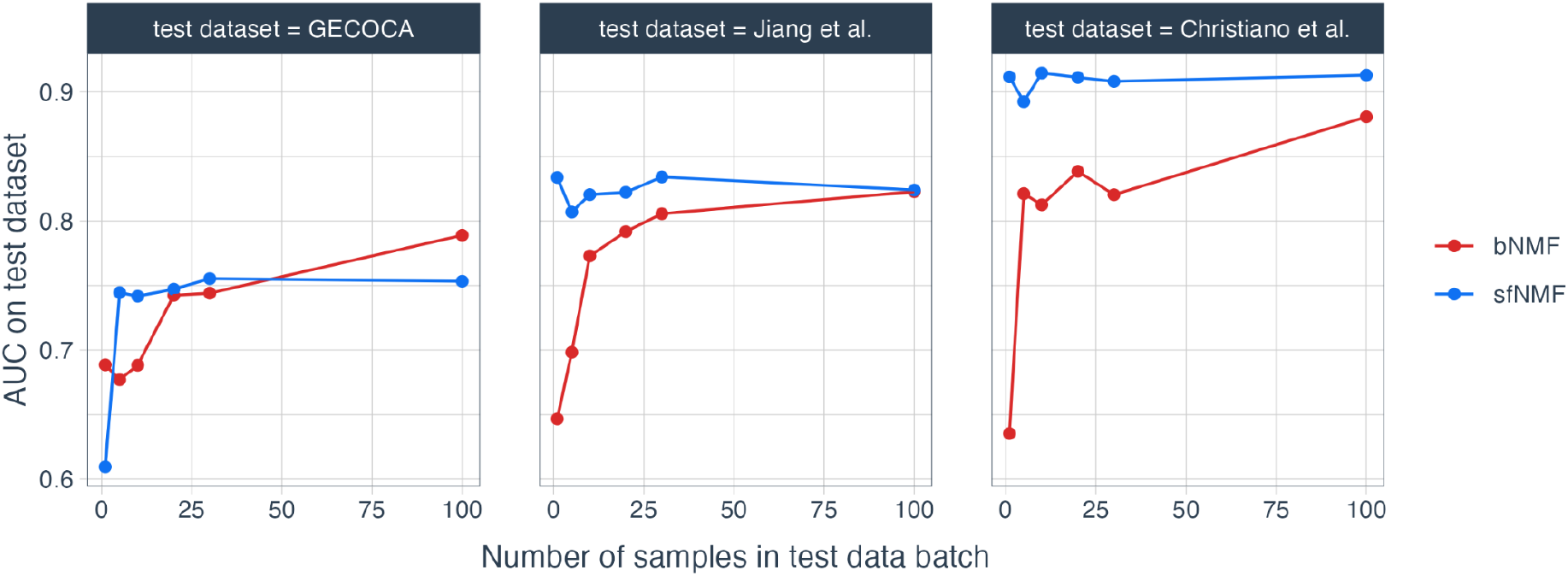
Results from leave-one-dataset-out cross-validation with 20 signatures when the test dataset is divided into smaller batches of 1, 5, 10, 20, 30, or 100 samples, and each of these smaller batches is analysed independently.

### 3.3 Models trained across all three datasets

We fitted each of the three NMF models with 20 signatures across all three datasets. To investigate the similarity of the signature weights in the control samples across different datasets, we performed a principal component analysis to determine if they clustered together (Fig. 8; Supp. Fig. 1). Using standard NMF, the control samples from each dataset formed distinct clusters when plotted on the first two principal components. For bNMF and sfNMF, the control samples do not form separate clusters and are much more similar. We also observed that if we divide the input matrix by the learned batch effect/filters matrix, the differences between the average distance distributions in the datasets get smaller (Supp. Fig. 2.).

**Figure 8.**
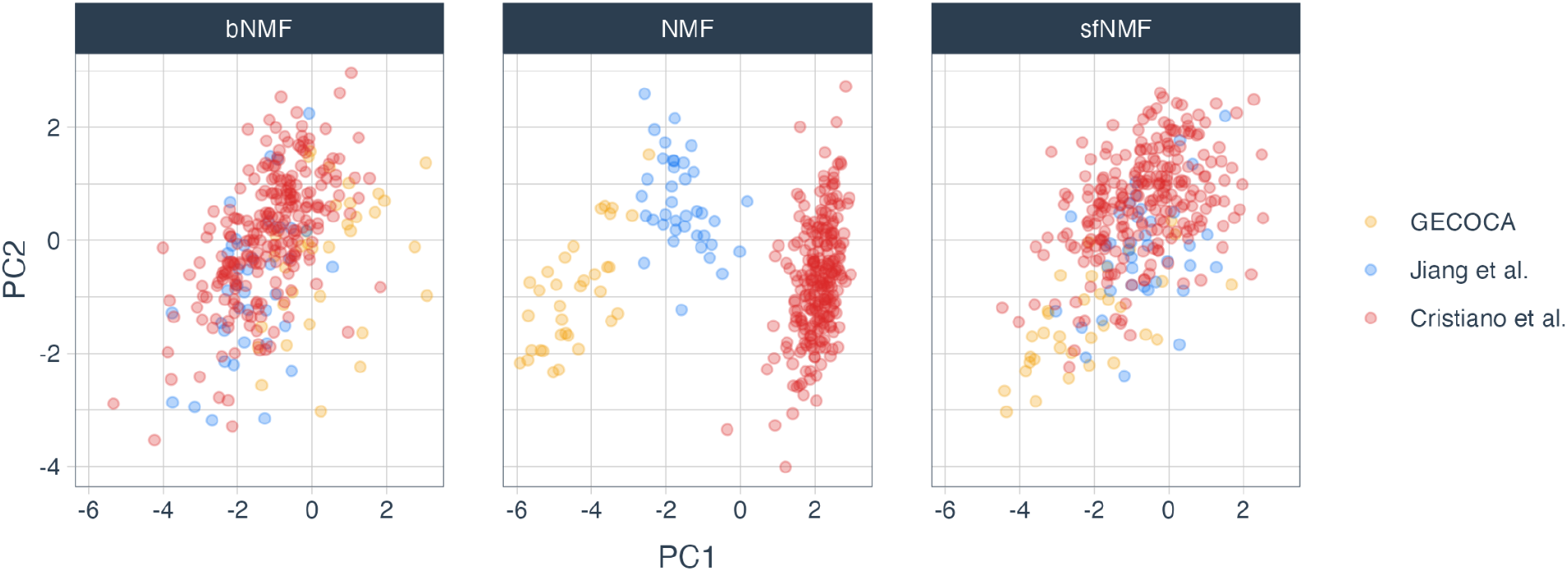
Results of a principal component analysis of the inferred signature weights. The plot shows the first two principal components for all controls across the three methods. Using standard NMF, the controls from the three different datasets can be easily separated, whereas the controls from different datasets appear more similar when using the two proposed NMF extensions.

In NMF models with two signatures, we obtain one signature that reflects tumor DNA and another that reflects cfDNA from healthy cells (Renaud et al. 2022). Models with a larger number of signatures fit the data better, but are more difficult to interpret. Figure 9 shows the inferred signatures, plotted with colors determined by the size of their LASSO coefficients. In both bNMF (Fig. 9a) and sfNMF (Fig. 9b), the signature that is the strongest indicator of cancer is enriched for small fragments less than 167bp. Both methods also exhibit cancer-associated signatures, with a peak at approximately 310 bp, and non-cancer-associated signatures, with peaks at around 330 bp. This suggests that short di-nucleosome fragments are indicative of cancer, while long di-nucleosome fragments are associated with healthy circulating DNA (cfDNA). The complete set of signatures annotated with the exact LASSO coefficients can be seen in Supp. Figures 4 and 5. Supp. Figure 7 shows the batch effects matrix and filter matrix that were fitted with bNMF and sfNMF, respectively.

**Figure 9.**
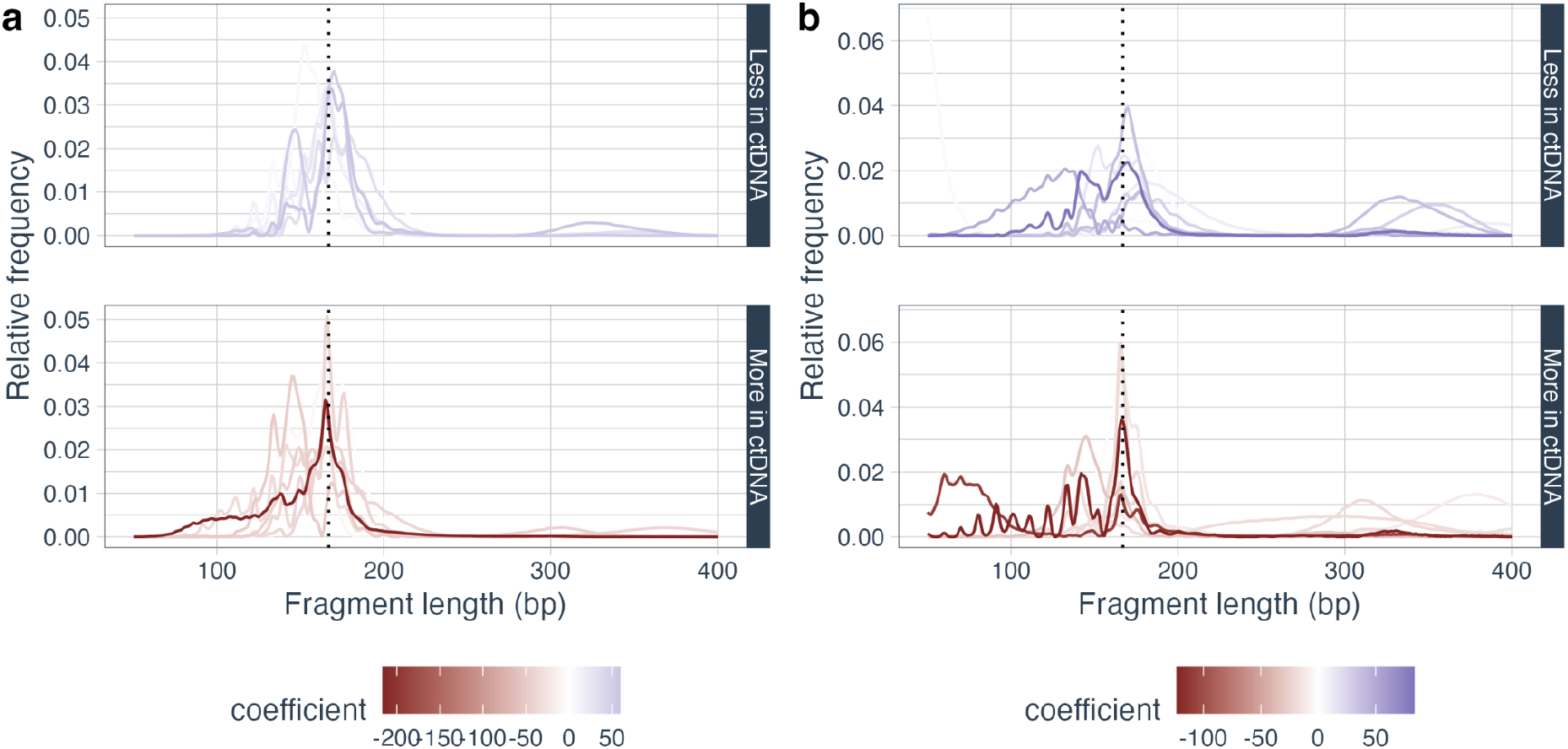
Used fragment length signatures. **a**. The signatures identified by bNMF are displayed with colors indicating their coefficients in the LASSO model. Signatures with a positive coefficient (enriched in healthy cfDNA) are in the top panel, and signatures with negative coefficients are in the bottom panel. **b**. The signatures found by sfNMF. The color indicates their coefficient in the LASSO model.

## 4 Conclusion and future directions

We have described two extensions of NMF that both address the problem of batch differences in data comprising multiple datasets with different technical biases. bNMF is a general solution to this problem that will also be useful for other types of data. It could, for instance, be used to model the fragment end-motifs on the same cfDNA data as used in this article, since these are affected by the same analytical and preanalytical biases as the fragment lengths. sfNMF is a model that assumes that the differences between datasets can be explained by a four-parameter continuous filter function. We use the knowledge that length *x* is followed by *x +* 1, and that the amount of filtering on *x +* 1 should be similar to the amount of filtering on *x* The sfNMF solution is thus specifically designed to work on fragment-length data. It can not be applied to other features, such as end-motifs, where there is no natural ordering of the motifs and no assumption of similarity between neighboring columns in the data.

We tested the methods on data comprising three datasets produced using different protocols in different labs. We found that both methods achieved approximately 10% improvements in AUC on independent test data compared to plain NMF, with bNMF performing slightly better than sfNMF. Although it does not outperform bNMF, sfNMF has the advantage of using significantly fewer parameters. This enables us to apply a model to a small test dataset with only a few samples. It is not possible to apply the bNMF model to a single test sample since the model would then need to estimate *C* + *M* parameters (in *W* and *E*) based on *M* data points (in *X*), and would thus be able to fit the data perfectly regardless of the signatures. This is not a problem for the sfNMF model, where we only need to estimate *C +* 4 parameters.

An issue with NMF is selecting the number of signatures to use. An improvement to the current work would be to incorporate an automatic strategy for selecting the optimal number of signatures based on the training data, for example, using a cross-validation strategy or an information criterion to do so. Typically, there can be several solutions to the NMF problem that achieve the same loss value. Recent articles have demonstrated that incorporating regularization based on the volume of the signature matrix can aid in selecting the optimal solution among them (Jin et al.,(Jin et al. 2024). Another improvement of the bNMF and sfNMF methods would be to add such regularisation.

Our implementation of bNMF and sfNMF in PyTorch utilizes transformations to ensure non-negativity and is very flexible, making it easy to modify. This way of fitting the models is, however, slow compared to fitting NMF using the multiplicative update rules. Another possible improvement to the current work would be to derive multiplicative update rules for the bNMF and sfNMF models as well, creating a faster method for fitting these models.

A general issue with cfDNA fragmentomics methods is that they are much more sensitive to technical differences between datasets than cfDNA methods that use tumor mutations or copy number alterations to detect cancer. This means that we, in general, cannot expect such methods to work well on datasets that have not been produced using the same lab protocols as the training data. It is a major limitation of such methods that they generally require retraining on data that matches the test data. In this study, we demonstrate that the transferability of a fragment-length classifier can be improved by utilizing one of our extended NMF methods to extract features. We can now create a trained model that we can expect to work on unseen datasets, even if they have different technical biases from the training data.

## Data Availability

The cfDNA data sets used in the article are from previously published studies. The data sets of Cristiano et al. and Jiang et al. are available at EGA (EGAD00001005339, EGAD00001005093). The GECOCA dataset is available at GenomeDK (https://genome.au.dk/library/GDK000015/).

## Code Availability

The relevant code for applying bNMF and sfNMF is available at: https://github.com/BesenbacherLab/batch-NMF. The *ctDNAtool* for extracting bin-wise fragment length counts is available at https://github.com/BesenbacherLab/ctDNAtool

## Acknowledgements

This study was funded by grants from Sygesikring Danmark (2020-0412), Aarhus University Research Foundation (AUFF-F-2020-7-8), and Novo Nordisk Fonden (NNF21OC0069056).

## Supplementary Material

**Supplementary Table 1:** Technical processing steps used on the different datasets.

| **Dataset** | **Sequencer** | **cfDNA Isolation** | **Library Prep** | **Amplification** | **Library cleanup / purification** |
| --- | --- | --- | --- | --- | --- |
| GECOCA | Illumina NovaSeq v1.5 | Qiagen QIAamp Circulating Nucleic Acids Kit | KAPA HyperPrep library kit | 7-cycle PCR | Cleanup: AMPure beads in a 1.4× beads/DNA ratio<br>Purification: AMPure beads in a 1.0× beads/DNA ratio |
| Cristiano et al. 2019 | Illumina HiSeq 2000/2500 | Qiagen Circulating Nucleic Acids Kit | NEBNext DNA Library Prep Kit | Phusion Hot Start Polymerase | Purification: AMPure XP beads |
| Jiang et al. 2015 | Illumina HiSeq 2000 | QIAamp DSP DNA Blood Mini Kit (Qiagen) | Kapa Library Preparation Kit | 14-cycle PCR (KAPA HiFi HotStart ReadyMix PCR Kit) | N/A |

**Supplementary Figure 1:**
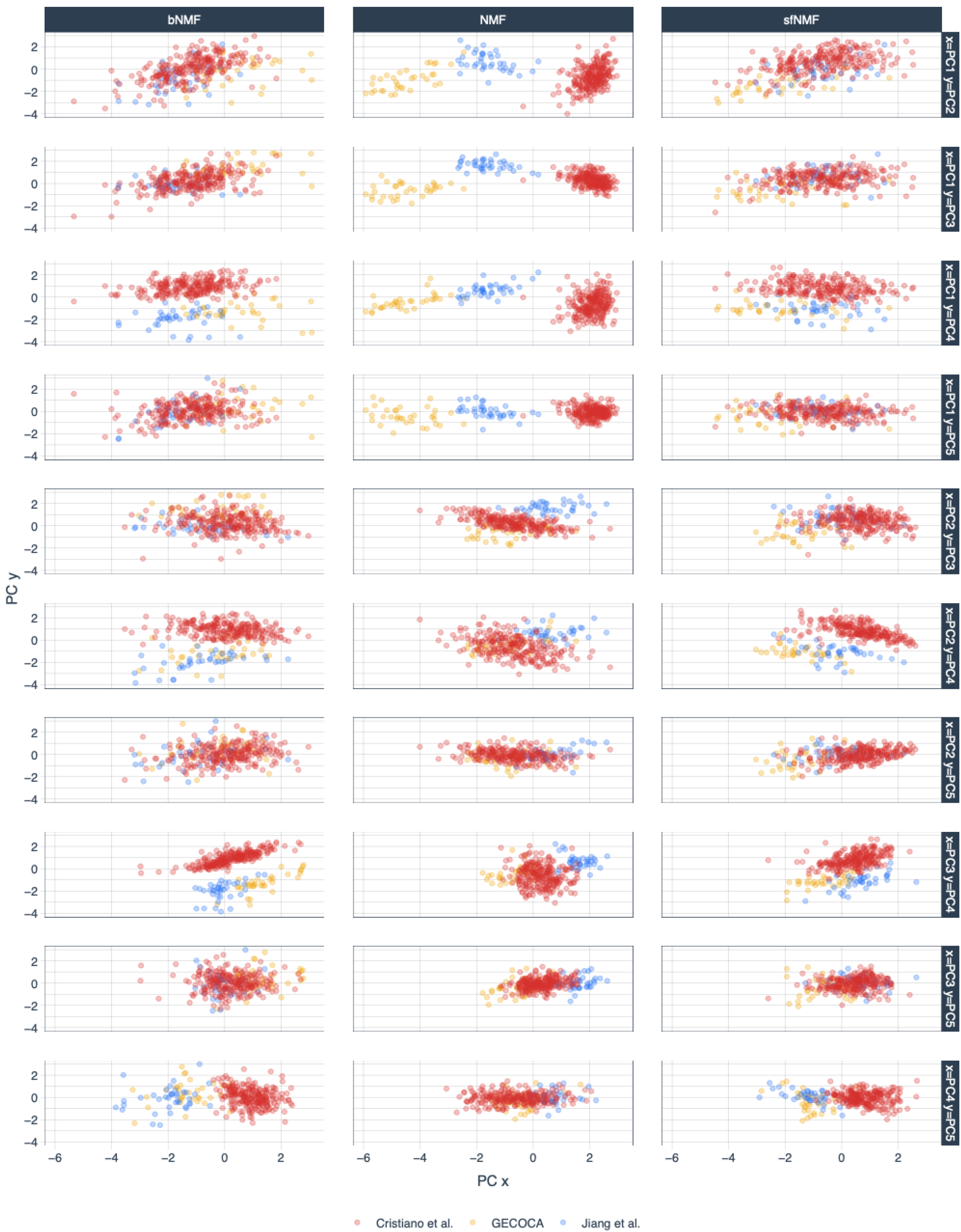
Separation of datasets (controls only) from combinations of the first five principal components of the NMF weights per NMF method.

**Supplementary Figure 2:**
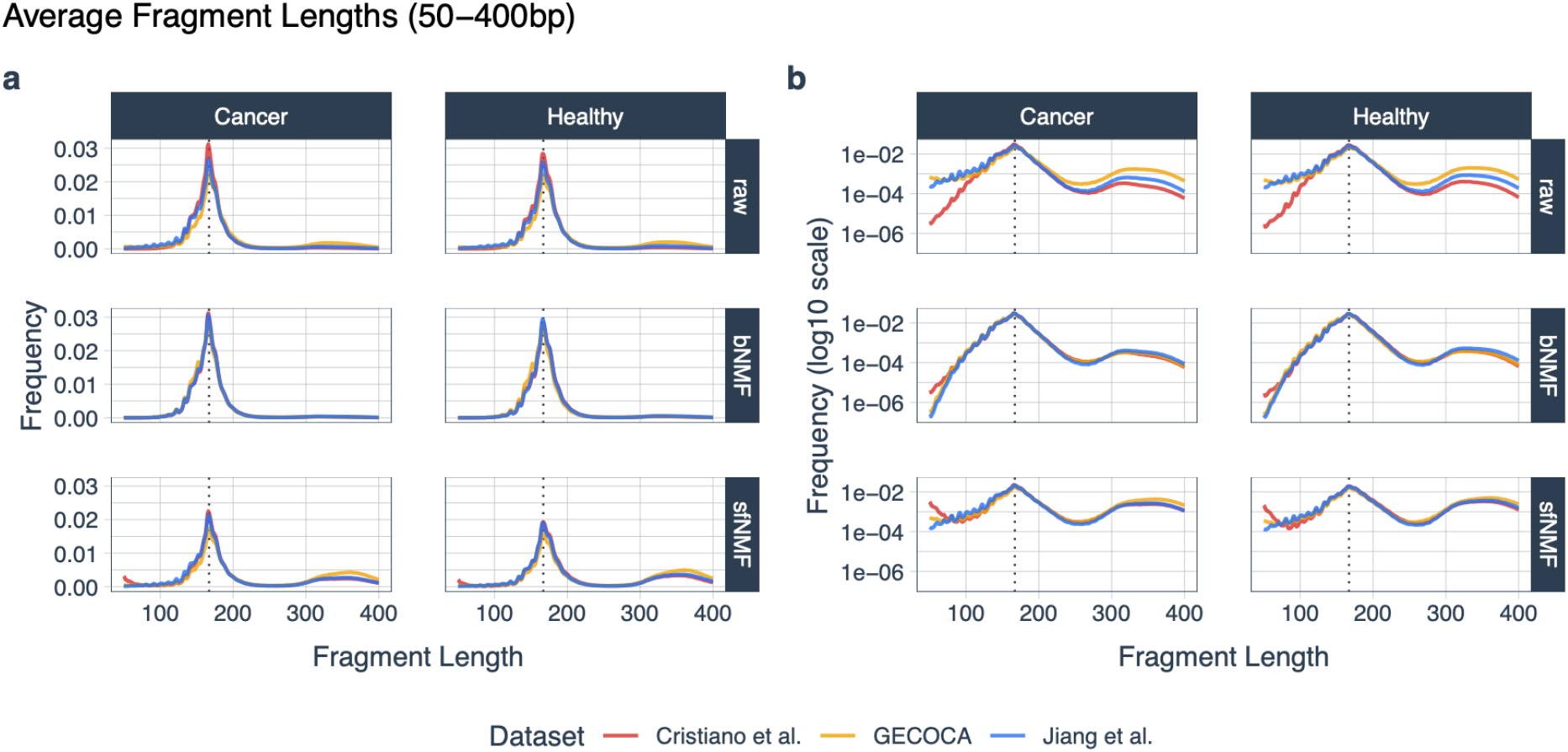
Average fragment length distributions per phenotype (horizontal panes), filtering method (vertical panes), and dataset (line color), visualizing the dataset differences. For the bNMF and sfNMF methods, the filters learned in the three-dataset model were corrected for in the counts via division (i.e. counts / filter) before any log transformation and sum-to-one normalization. **a**. Frequency of the fragment lengths. **b**. Same as **a** but on a log-10 scale.

**Supplementary Figure 3:**
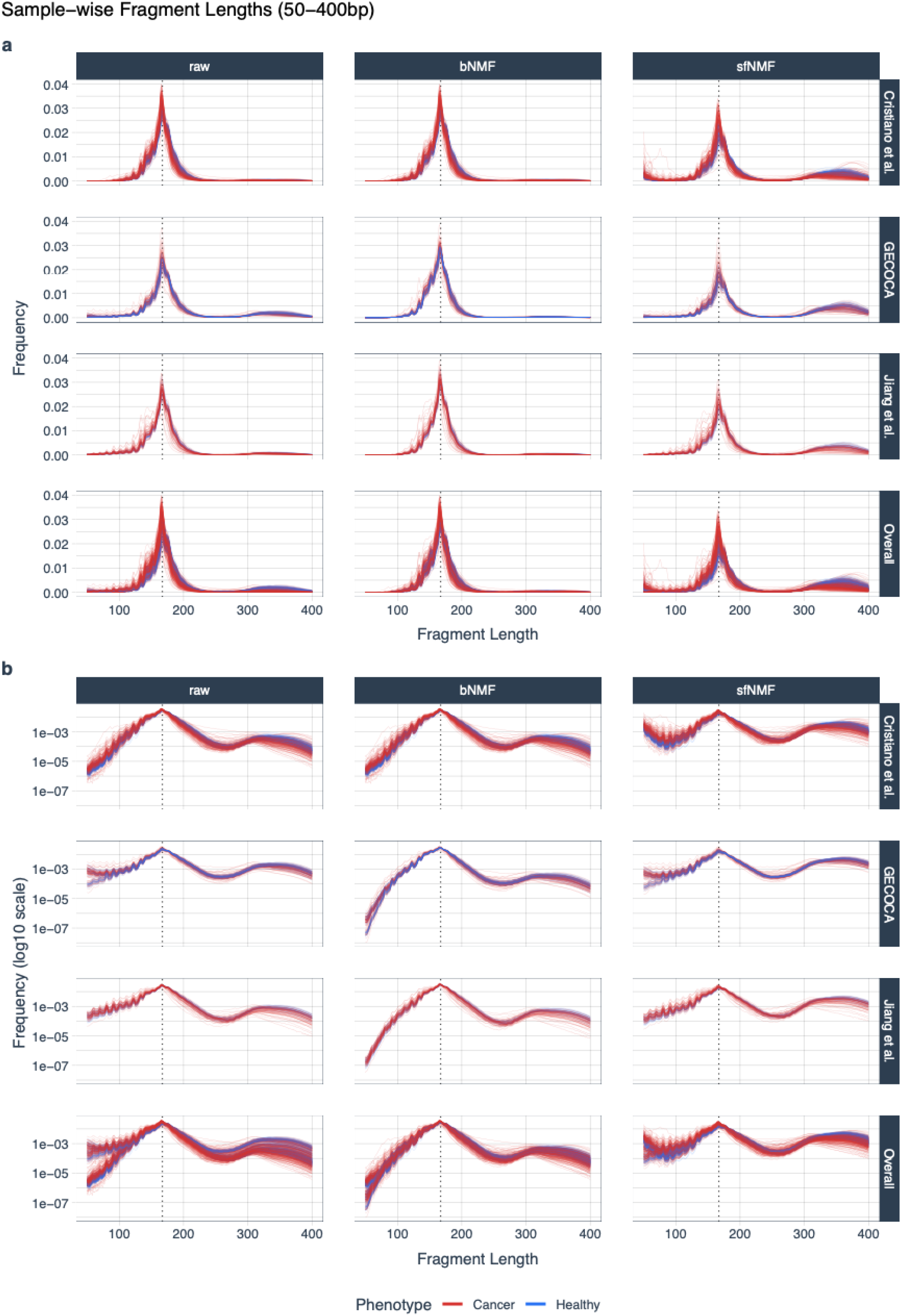
Sample-wise fragment length distributions per filtering method (horizontal panes), dataset (vertical panes) and phenotype (line color), visualizing the control-cancer differences. For the bNMF and sfNMF methods, the filters learned in the three-dataset model were corrected for in the counts via division (i.e. counts / filter) before any log transformation and sum-to-one normalization. The “Overall” row contains the overall average curves. **a**. Frequency of the fragment lengths. **b**. Same as **a** but on a log-10 scale.

**Supplementary Figure 4:**
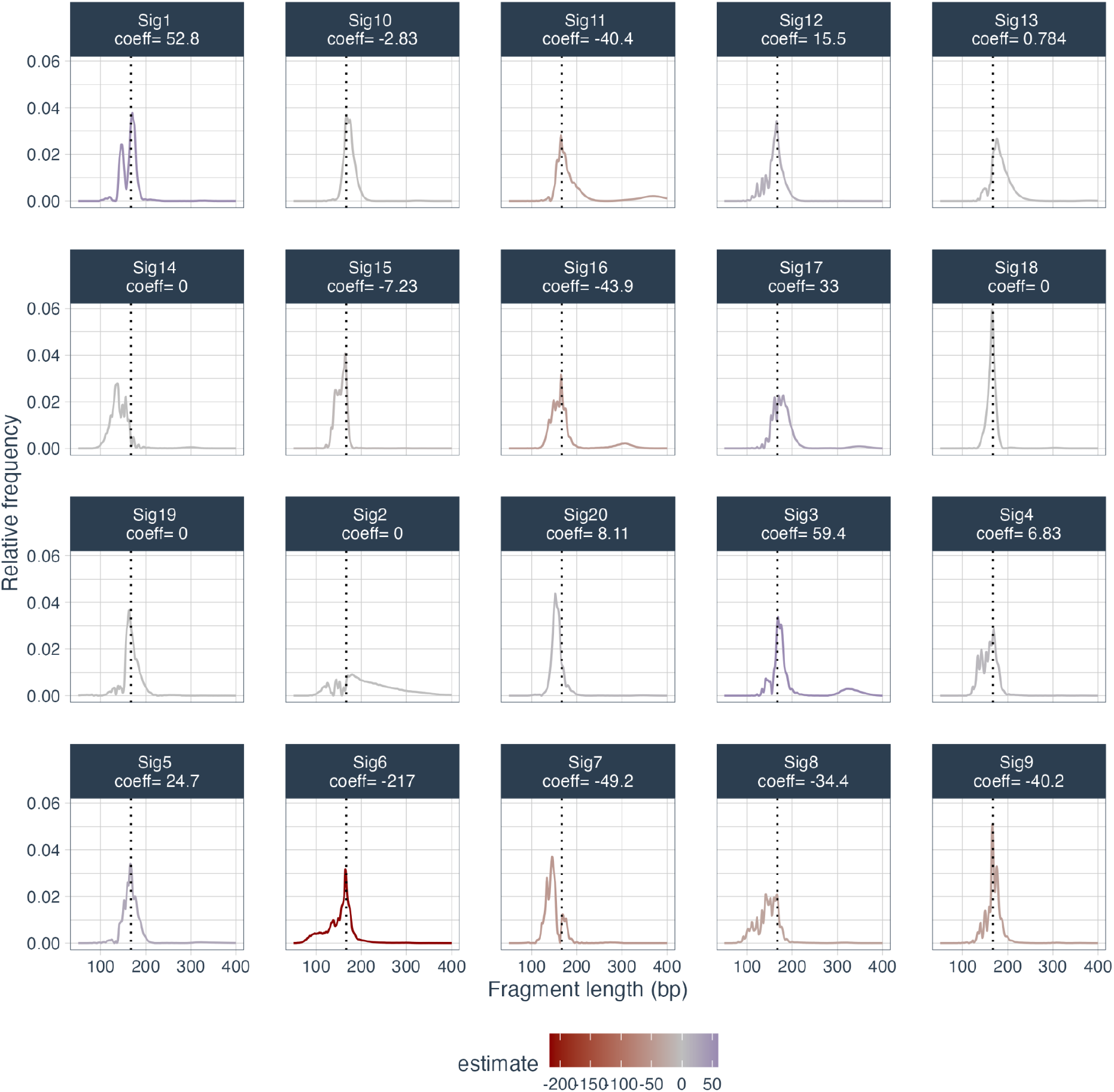
The 20 signatures estimated by bNMF when run on the combined dataset. Their coefficients estimated in the LASSO model are listed under their names, and the color of the line corresponds to each one. A lower coefficient indicates that the signature is more prevalent in cancer samples.

**Supplementary Figure 5:**
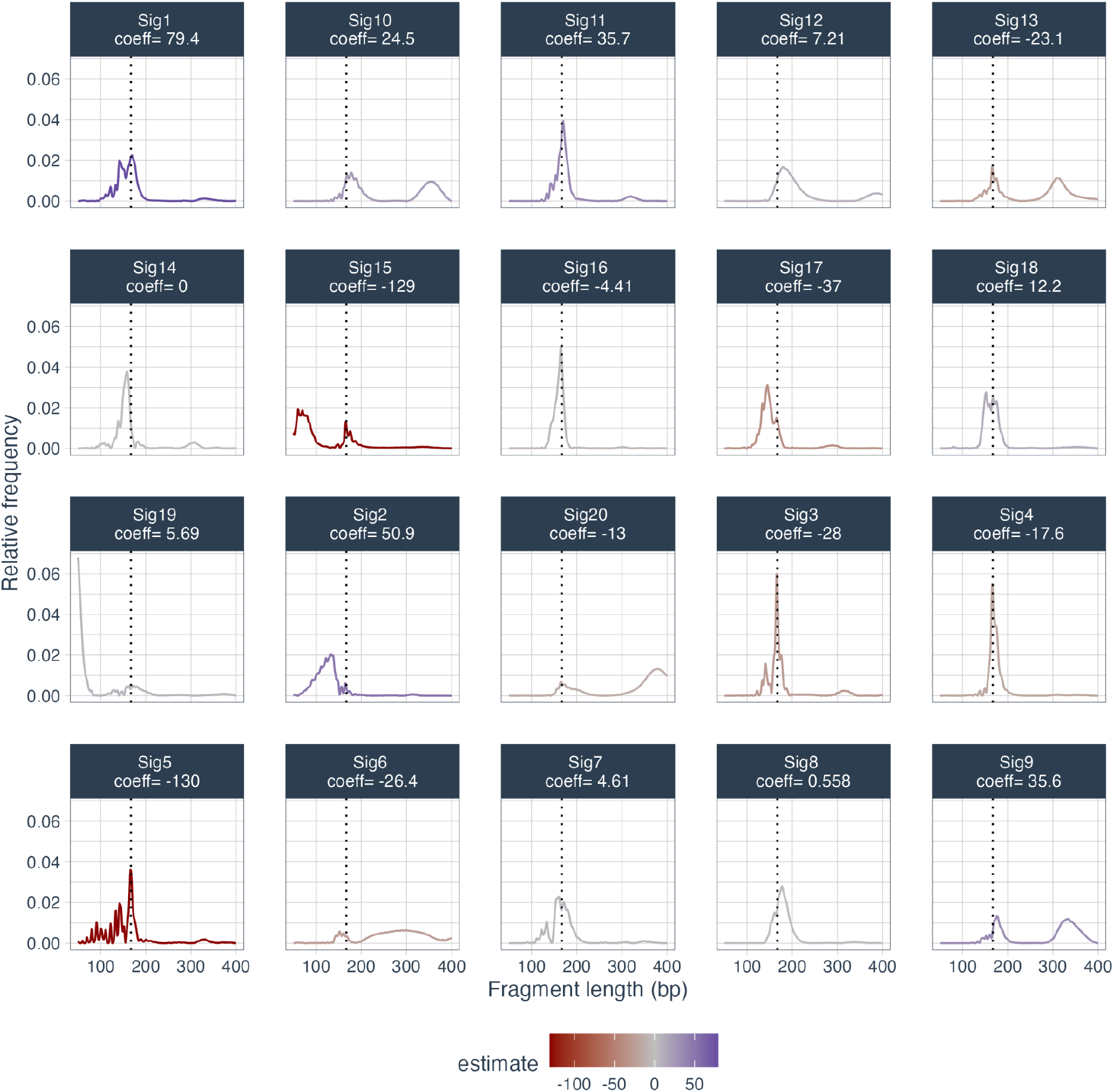
The 20 signatures estimated by sfNMF when run on the combined dataset. Their coefficients estimated in the LASSO model are listed under their names, and the color of the line reflects it. A lower coefficient indicates that the signature is more prevalent in cancer samples.

**Supplementary Figure 6:**
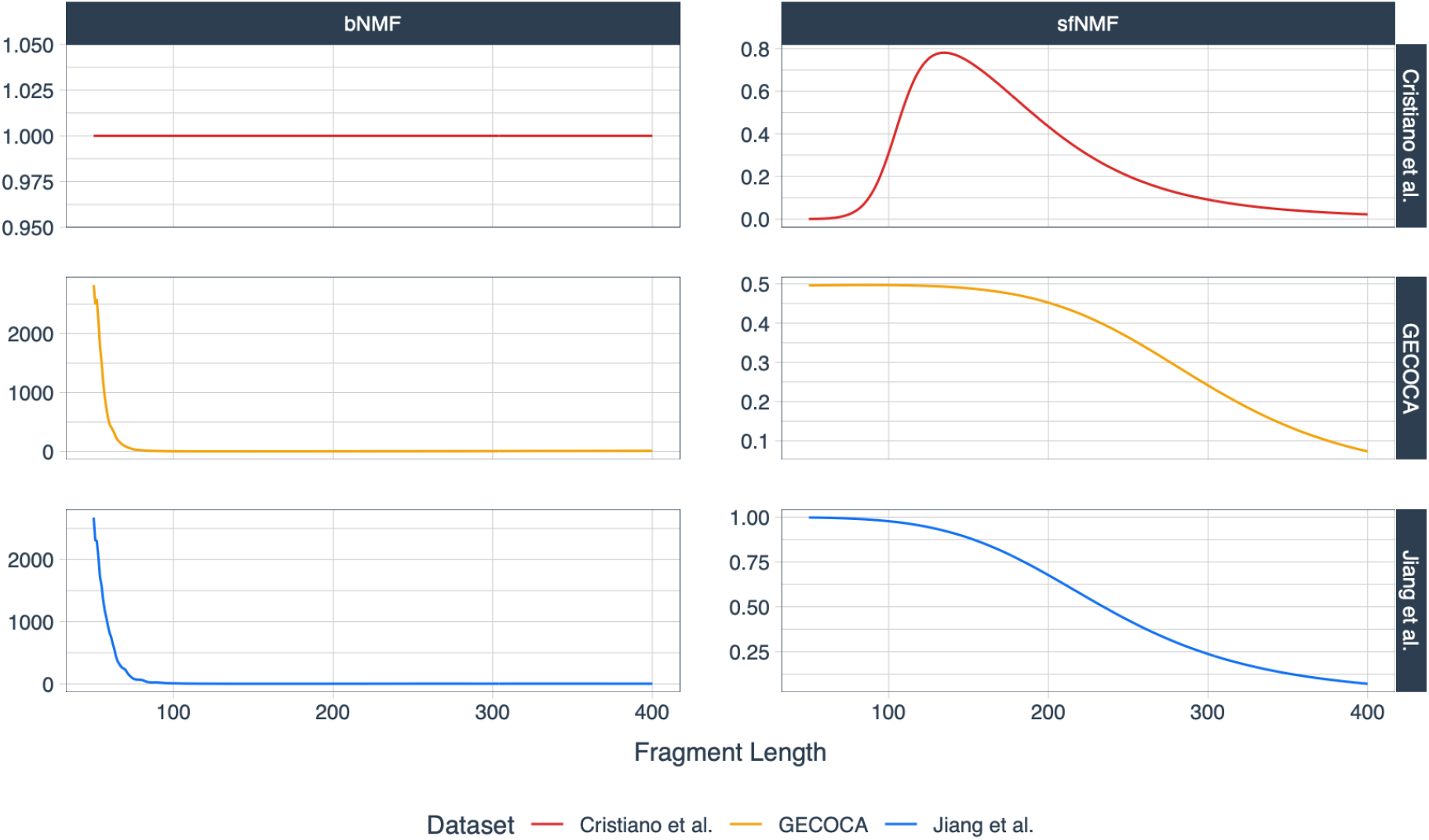
The fitted batch effects / filters. The left panel shows the fitted batch effect matrix, *E*, when running bNMF with 20 signatures. The Cristiano et al. (top panel) was the reference dataset, and the effects in the two other datasets are relative to that dataset. The right panel shows the fitted filters when running sfNMF with 20 signatures.

## References

Alexandrov, Ludmil B., Serena Nik-Zainal, David C. Wedge, Peter J. Campbell, and Michael R. Stratton. 2013. “Deciphering Signatures of Mutational Processes Operative in Human Cancer.” Cell Reports 3 (1): 246–59.

Amemiya, Haley M., Anshul Kundaje, and Alan P. Boyle. 2019. “The ENCODE Blacklist: Identification of Problematic Regions of the Genome.” Scientific Reports 9 (1): 9354.

Bai, Jinyue, Peiyong Jiang, Lu Ji, W. K. Jacky Lam, Qing Zhou, Mary-Jane L. Ma, Spencer C. Ding, et al. 2024. “Histone Modifications of Circulating Nucleosomes Are Associated with Changes in Cell-Free DNA Fragmentation Patterns.” Proceedings of the National Academy of Sciences of the United States of America 121 (42): e2404058121.

Chang, Adrienne, Omary Mzava, Joan S. Lenz, Alexandre P. Cheng, Philip Burnham, S. Timothy Motley, Crissa Bennett, et al. 2021. “Measurement Biases Distort Cell-Free DNA Fragmentation Profiles and Define the Sensitivity of Metagenomic Cell-Free DNA Sequencing Assays.” Clinical Chemistry 68 (1): 163–71.

Cristiano, Stephen, Alessandro Leal, Jillian Phallen, Jacob Fiksel, Vilmos Adleff, Daniel C. Bruhm, Sarah Østrup Jensen, et al. 2019. “Genome-Wide Cell-Free DNA Fragmentation in Patients with Cancer.” Nature 570 (7761): 385–89.

Dabney, Jesse, and Matthias Meyer. 2012. “Length and GC-Biases during Sequencing Library Amplification: A Comparison of Various Polymerase-Buffer Systems with Ancient and Modern DNA Sequencing Libraries.” BioTechniques 52 (2): 87–94.

Giacona, M. B., G. C. Ruben, K. A. Iczkowski, T. B. Roos, D. M. Porter, and G. D. Sorenson. 1998. “Cell-Free DNA in Human Blood Plasma: Length Measurements in Patients with Pancreatic Cancer and Healthy Controls.” Pancreas 17 (1): 89–97.

Gohl, Daryl M., Alessandro Magli, John Garbe, Aaron Becker, Darrell M. Johnson, Shea Anderson, Benjamin Auch, et al. 2019. “Measuring Sequencer Size Bias Using REcount: A Novel Method for Highly Accurate Illumina Sequencing-Based Quantification.” Genome Biology 20 (1): 85.

Han, Diana S. C., Meng Ni, Rebecca W. Y. Chan, Vicken W. H. Chan, Kathy O. Lui, Rossa W. K. Chiu, and Y. M. Dennis Lo. 2020. “The Biology of Cell-Free DNA Fragmentation and the Roles of DNASE1, DNASE1L3, and DFFB.” The American Journal of Human Genetics 106 (2): 202–14.

Jahr, S., H. Hentze, S. Englisch, Dieter Hardt, F. O. Fackelmayer, R. Hesch, and R. Knippers. 2001. “DNA Fragments in the Blood Plasma of Cancer Patients: Quantitations and Evidence for Their Origin from Apoptotic and Necrotic Cells.” Cancer Research 61 (4): 1659–65.

Jiang, Peiyong, Carol W. M. Chan, K. C. Allen Chan, Suk Hang Cheng, John Wong, Vincent Wai-Sun Wong, Grace L. H. Wong, et al. 2015. “Lengthening and Shortening of Plasma DNA in Hepatocellular Carcinoma Patients.” Proceedings of the National Academy of Sciences of the United States of America 112 (11): E1317–25.

Jin, Hu, Doga C. Gulhan, Benedikt Geiger, Daniel Ben-Isvy, David Geng, Viktor Ljungström, and Peter J. Park. 2024. “Accurate and Sensitive Mutational Signature Analysis with MuSiCal.” Nature Genetics, February. 10.1038/s41588-024-01659-0.

Karimzadeh, Mehran, Carl Ernst, Anshul Kundaje, and Michael M. Hoffman. 2018. “Umap and Bismap: Quantifying Genome and Methylome Mappability.” Nucleic Acids Research 46 (20): e120.

Lee, Daniel, and H. Sebastian Seung. 2000. “Algorithms for Non-Negative Matrix Factorization.” Advances in Neural Information Processing Systems 13. https://proceedings.neurips.cc/paper_files/paper/2000/file/f9d1152547c0bde01830b7e8bd60024c-Paper.pdf.

Markus, Havell, Tania Contente-Cuomo, Maria Farooq, Winnie S. Liang, Mitesh J. Borad, Shivan Sivakumar, Simon Gollins, et al. 2018. “Evaluation of Pre-Analytical Factors Affecting Plasma DNA Analysis.” Scientific Reports 8 (1): 7375.

Mouliere, Florent, Dineika Chandrananda, Anna M. Piskorz, Elizabeth K. Moore, James Morris, Lise Barlebo Ahlborn, Richard Mair, et al. 2018. “Enhanced Detection of Circulating Tumor DNA by Fragment Size Analysis.” Science Translational Medicine 10 (466). 10.1126/scitranslmed.aat4921.

Olsen, Ludvig Renbo, Denis Odinokov, Jakob Qvortrup Holsting, Karoline Kondrup, Laura Iisager, Maria Rusan, Simon Buus, et al. 2024. “Cross-Dataset Pan-Cancer Detection: Correlating Cell-Free DNA Fragment Coverage with Open Chromatin Sites across Cell Types.” medRxiv. 10.1101/2024.11.26.24317971.

Renaud, Gabriel, Maibritt Nørgaard, Johan Lindberg, Henrik Grönberg, Bram De Laere, Jørgen Bjerggaard Jensen, Michael Borre, et al. 2022. “Unsupervised Detection of Fragment Length Signatures of Circulating Tumor DNA Using Non-Negative Matrix Factorization.” eLife 11 (July). 10.7554/eLife.71569.

Rodrigue, Sébastien, Arne C. Materna, Sonia C. Timberlake, Matthew C. Blackburn, Rex R. Malmstrom, Eric J. Alm, and Sallie W. Chisholm. 2010. “Unlocking Short Read Sequencing for Metagenomics.” PloS One 5 (7): e11840.

Serpas, Lee, Rebecca W. Y. Chan, Peiyong Jiang, Meng Ni, Kun Sun, Ali Rashidfarrokhi, Chetna Soni, et al. 2019. “Dnase1l3 Deletion Causes Aberrations in Length and End-Motif Frequencies in Plasma DNA.” Proceedings of the National Academy of Sciences of the United States of America 116 (2): 641–49.

Shi, Jiping, Runling Zhang, Jinming Li, and Rui Zhang. 2020. “Size Profile of Cell-Free DNA: A Beacon Guiding the Practice and Innovation of Clinical Testing.” Theranostics 10 (11): 4737–48.

Thierry, A. R. 2023. “Circulating DNA Fragmentomics and Cancer Screening.” Cell Genomics 3 (1): 100242.

Wang, Brant G., Han-Yao Huang, Yu-Chi Chen, Robert E. Bristow, Keyanunoosh Kassauei, Chih-Chien Cheng, Richard Roden, Lori J. Sokoll, Daniel W. Chan, and Ie-Ming Shih. 2003. “Increased Plasma DNA Integrity in Cancer Patients.” Cancer Research 63 (14): 3966–68.

Wang, Haichao, Paulius D. Mennea, Yu Kiu Elkie Chan, Zhao Cheng, Maria C. Neofytou, Arif Anwer Surani, Aadhitthya Vijayaraghavan, et al. 2025. “A Standardized Framework for Robust Fragmentomic Feature Extraction from Cell-Free DNA Sequencing Data.” Genome Biology 26 (1): 141.

Zhou, Qing, Guannan Kang, Peiyong Jiang, Rong Qiao, W. K. Jacky Lam, Stephanie C. Y. Yu, Mary-Jane L. Ma, et al. 2022. “Epigenetic Analysis of Cell-Free DNA by Fragmentomic Profiling.” Proceedings of the National Academy of Sciences of the United States of America 119 (44): e2209852119.

